# Determinants of contraceptive use and intention to use among youth 15-24 years in Karamoja, Uganda

**DOI:** 10.1101/2025.01.09.25320305

**Authors:** Lillian Ojanduru, Godfrey Siu, Justine Bukenya, Nazarius. M. Tumwesigye

**Author notes:** **Corresponding author**: Lillian Ojanduru: Makerere University School of Public Health, **Email:****, **.

## Abstract

**Background:** Globally, contraceptive use among young people aged 15-24 years is low with a prevalence lower than 21% yet contraceptive use reduces the likelihood of high-risk and unintended pregnancies. In Uganda, nearly a quarter (24%) of girls aged 15-19 years have either been pregnant or given birth, mainly attributed to non-use of contraceptives which is so low at 10% in Karamoja. In response the study aimed to assess determinants of contraceptive use and intention to use in Karamoja region

**Methods:** This was a quantitative study with a sample size of 448 youth (women and men 15-24 years). Of these,409 were sexually active (had sex within the last 12 months) and included in our analysis. Modified Poisson model that incorporated sampling weights was used to establish the factors associated with contraceptive use, and intention to use.

**Results:** About 41.6% (49.3% males and 32.6% females) had ever used a contraceptive. only 11.0% (16.3% women and 6.4% men) were currently using a contraceptive and about 72.4% (84% males and 59% females) intended to use contraception. Youths with secondary education had higher prevalence rate for ever use of contraceptives by 32% and 25% for females and males respectively than those with no education. Females who had a paid job were 4.5 times (APR=4.51, 95%CI: 3.80-5.36) and males were 1.6 times (APR=1.6, 95% CI: 1.26-1.92) more likely to use contraceptives while the prevalence of intention to use was higher among those who had a job by 5.7 times (APR=5.75, 95%CI: 4.94-6.69) for females and 2.3 times (APR=2.25, 95%CI: 1.76-2.89) for males. Never married females were 3.1 times (APR=3.09, 95%CI: 2.39-3.99) more likely to use contraceptives than married females. Females who had children 1.6 times (APR=1.46, 95% CI: 1.18-1.81) and males 7.4 times (APR=7.4, 95% CI: 4.74-11.69) more like to have ever used a contraceptive.

**Conclusion:** The factors positively associated with use of contraceptives and intention to use were age (being older, 20-24 year), marital status, having biological children, attaining secondary education level, living with biological parents and having a job and all these varied by gender. The study recommends improving access to education and employment opportunities for youth, involving parents in young people’s SRH, and targeting male youth with interventions to improve contraceptive use.

## 1. Introduction

Every year, an estimated 21 million girls aged 15–19 years in developing regions become pregnant and approximately 12 million of them give birth of which 50% are unintended (1). In Africa, 20% of all pregnancies occur in adolescents, with a big burden in the East African Region (2) yet their sexual and reproductive health (SRH) needs have remained largely unmet. In 2021, the estimated number of births among 15–19-year-olds in Sub-Saharan Africa (SSA) was 6,114,000 and the rate of adolescent fertility in SSA was higher than the world average, with 100 out of every 1,000 girls aged 15–19 giving birth (3). Many of births are unintended and attributed to low contraceptive use (1). In 2019, globally 10.2% of sexually active adolescent women 15 - 19 were using modern contraceptives with low prevalence and high unmet need in Sub-Saharan Africa (4). Uganda has one of the highest rates of teenage pregnancy in sub-Saharan Africa. Nearly a quarter (24%) of girls aged 15 to 19 years have either been pregnant or given birth (5, 6), with most pregnancies taking place within marriage, reflecting a high level (49%) of child marriage by their 18th birthday (UDHS, 2022). Uganda has one of the highest maternal mortality rates (189/100,000) live births with pregnancy-related mortality ratio of 228/1000,000 (5) and the highest among adolescents caused by complications from pregnancy and childbirth.

Uptake of contraceptives in Uganda among youth is generally low with only 9.4% female adolescents aged 15-19, and 33.4% of those aged 20–24 years using a method (5). Unmet need for family planning is high at 28.8% for youth aged 15-19 years and 22.7% for youth aged 20-24 years (5). However, unmet need for FP in Karamoja is 24.3% but both demand for family planning (43%) and use (10%) of modern contraceptives are lowest in Karamoja region (5) way below the national average of 38% of contraceptive use.

Many studies on contraceptive use tend to focus mainly on adolescent girls and women. Y15-24 years old, particularly males, are rarely included and this undermines the strategic efforts to improve contraceptive utilization given Uganda’s population is mainly young with 77% under the age of 25 (5) thus a greater need for increased contraceptive uptake This study addresses this gap by assessing determinants of contraceptive use and intention to use among both female and male youths.

## 2. Methodology

### 2.1 Study design and study site

We conducted a cross-sectional study and collected quantitative data in four districts of Karamoja-Moroto, Nakapiripirit, Napak and Nabilatuk. The data collection took place from the 25^th of^ March 2023 to 15^th^ August 2023.

### 2.2 Sampling and sample size

Using Taro Yamane’s sample size calculation formula (1967), we included a sample of 400 youths, at 90% power. Allowing for a 10% non-response at a precision of 0.05 (7), the adjusted sample was 448. However, the total number of sexually active youth (those who reported having intercourse in the last 12 months) interviewed and included in the analysis were 409 (190 women and 219 men).

Sampling: Simple random sampling was used to select both the study areas and households. Four parishes, from two sub counties in two districts were selected and all villages in the selected parishes were included in the study. Only one youth was interviewed per household. Data collection: Data were collected using mobile device using kobo collect. Data were uploaded to a central server where the supervisor checked for consistency and completeness.

### 2.3 Data analysis

Survey data from the central server were imported into Stata version 13 for analysis. Percentages were used to present the demographic and socio-economic characteristics of youth. Modified Poisson regression model was used because it is preferred in cross-sectional studies when the outcome of interest is not rare, and it approximates the risk ratios or relative ratios better than binary logistic regression model (8). In the case of our study, the outcome variable contraceptive use and intention to use contraceptives is not rare. The link function of modified Poisson regression model is log link.

The outcome variables were Contraceptive use and intention to use. Contraceptive use was measured by current and ever use of contraceptives and intention to use was measured by weather the research participants were intending to use contraceptives in the future.

### 2.4 Variables

The independent variables were;-age, sex, marital status, education level, listening to a radio, type of job having children, whom the youth live with and religion. We analyzed contraceptive use, and intention to use FP among sexually active youth and included these variables in one model to get adjusted prevalence ratios. We ran both unadjusted and adjusted for all variables except for religion because at the unadjusted level for female youth, there was no significant relationship between religion and intention to use contraceptives.

### 2.6 Ethics statement

This study was conducted in accordance with the Helsinki Declaration. This was part of a PhD study that was approved by the Makerere University School of Public Health, Research Ethics Committee (MUSPH-REC), approval number (SPH-2022-294). The study was also approved by Uganda National Council of Science and Technology (UNSCT), a national approving body (Internal Review Board), The UNCST approval number for the study is HS2547ES. Districts where the study was conducted provided letters of approval. All respondents who participated in the study provided a written informed consent except for parents or guardians of minors under the age of 18 who provided verbal informed consent. All consents were provided before the interview could start. The written consent was obtained in local language and in the presence of a witness for those unable to read and write. The witness was either a friend or community leader who could read and write to ensure all information on the consent form was read to the responded. The witness was not part of the study participant. All consent forms were signed and thump prints for those unable to write. The consent form made it clear that participation in the study was voluntary. Respondents were left with a copy of the consent form that had names and contact numbers of both the chair of the Research Ethics Committee and the Principal investigator in case if they had any questions about the study and or their rights as study participants and another copy for the research team. Respondents were assured of safety, confidentiality, voluntary participation and withdraw from the survey without any penalty. For participants who were younger than the age of 18 years, informed consent was obtained from their parents or guardians.

## Results

### 3.1 Characteristics of Respondents

Table 1 shows the characteristics of the respondents by socio-demographic factors. Out of the 448 participants interviewed, 91.3% (98% males and 84% females) were sexually active (had sex in the past 12 months). The majority (84.8%) were aged 20 to 24 years, and slightly more than half (57.2%) were married (44.8% males Vs 71.6% females). Overall, most of the participants had children (62.4%), although more females (84.7%) than males (42.9%) had children. While 35.0% (43.5% males Vs 25.3% females) were living with their biological parents, a good percentage either lived on their own (28.6%) or with their husband/wife (26.6%)

**Table 1.** Socio-demographic characteristics of young people aged 15 to 24 years by sex.

| Background Characteristics | All (n=409) | Men (n=219) | Women(n=190) | P-value |
| --- | --- | --- | --- | --- |
| Age (years) |  |  |  |  |
| 15-19 | 62 (15.2) | 43(19.6) | 19 (10.0) | 0.007 |
| 20-24 | 347 (84.8) | 176 (80.4) | 171 (90.0) |  |
| Currently in a relationship |  |  |  |  |
| Yes | 372 (91) | 194 (88.6) | 178 (89.7) | 0.073 |
| No | 37 (0.9) | 25 (11.4) | 12 (6.3) |  |
| Marital Status |  |  |  |  |
| Married or living together. | 234 (57.2) | 98 (44.8) | 136 (71.6) | 0.000 |
| Separated/divorced. | 20 (4.9) | 7 (3.2) | 13 (6.8) |  |
| Never Married | 155 (37.9) | 114 (52.0) | 41 (21.6) |  |
| Have Children |  |  |  |  |
| Yes | 225 (62.4) | 94 (42.9) | 161 (84.7) | 0.000 |
| No | 154 (37.6) | 125 (27.1) | 29 (15.3) |  |
| Live with |  |  |  |  |
| Biological parents | 143 (35.0) | 95 (43.4) | 48 (25.3) | 0.001 |
| Guardians | 32 (7.8) | 27 (12.3) | 5 (2.6) |  |
| Older siblings | 8 (2.0) | 8 (3.6) | 0 (0.0) |  |
| Own | 117 (28.6) | 86 (39.3) | 31 (16.3) |  |
| Husband/wife | 109 (26.6) | 3 (1.4) | 106 (55.8) |  |
| Education Level |  |  |  |  |
| No education | 155 (28.1) | 52 (23.7) | 63 (33.2) | 0.069 |
| Primary | 214 (52.3) | 118(53.9) | 96 (50.5) |  |
| Secondary + | 80 (19.6) | 49 (22.4) | 31 (16.3) |  |
| Have a Job |  |  |  |  |
| Yes | 168 (41.1) | 64 (29.2) | 104 (54.7) | 0.000 |
| No | 241 (58.9) | 155 (70.8) | 86 (45.5) |  |
| Type of Job |  |  |  |  |
| No Job | 241 (58.9) | 155 (70.8) | 86 (45.3) |  |
| Farming/agriculture | 73 (17.8) | 29 (13.2) | 44 (23.2) |  |
| Other | 95 (23.3) | 35 (16.0) | 60 (31.5) | 0.000 |
| **Religion** |  |  |  |  |
| Anglican | 118 (28.9) | 44 (20.1) | 74 (38.9) |  |
| Catholic | 261 (63.8) | 163 (74.4) | 98 (51.6) |  |
| Moslem | 23 (5.6) | 11 (5.0) | 12 (6.3) | 0,000 |
| Others | 7 (1.7) | 1(0.5) | 6(3.1) |  |
| **Listening to a radio** |  |  |  |  |
| Almost everyday | 147 (35.9) | 82 (37.5) | 65 (34.2) |  |
| Once a week | 128 (31.3) | 88 (40.2) | 40 (21.0) | 0.000 |
| Less than once a week | 24 (5.9) | 6 (2.7) | 18 (9.5) |  |
| Not at all | 110 (26.9) | 43 (19.6) | 67 (15.3) |  |
| **Ever use contraceptives** |  |  |  |  |
| Yes | 170 (41.6) | 108 (49.3) | 62 (32.6) | 0.001 |
| No | 224 (54.8) | 97 (44.3) | 127 (66.9) |  |
| Don't know | 15 (43.7) | 14 (6.4) | 1(0.5) |  |
| **Current contraceptive Use** |  |  |  |  |
| Use | 45 (11.0) | 14 (6.4) | 31 (16.3) | 0.001 |
| Non-Use | 364(89.0) | 205 (93.6) | 159 (83.7) |  |
| **Intention to use contraceptives** |  |  |  |  |
| Yes | 296 (72 .4) | 184 (84.0) | 112 (59.0) | 0.000 |
| No | 113 (27.4) | 35 (16.0) | 78 (41.0) |  |

About 52.3% had attained primary school (53.9% males vs 50.4% females) and more than half (58.9%) did not have a job (70.8% males and 45.5% females) Slightly more than a third (35.9%) reported listening to radio almost every day (37.5% males and 34.2%females).

A total of 41.6% (49.3% males and 32.6% females) reported to have ever used or their partners had ever used contraceptives. This was highest among men aged 20-24 (54.0%), and women aged 20-24 (35.1%). However, only 11.0% (6.4% males and 16.3% females) were currently using a method and 72.4% (84.0% males and 59.0% females) had intention to use contraceptives.

### 3.2 Current use of contraceptives

Among female youth, about 16.3% (31/190) were currently using some form of contraceptive. Injectables were the most used method among 20-24-year-olds, reported by 58.1%, followed by implants at 41.9% and male condoms by 12.9% but use of other methods like pills, IUD, female condoms was extremely low, and no adolescent girls aged 15-19 years used any other method apart from injectables.

Among male youths, about 6.4% (14/219) were using a method of FP. Male condoms were the most used method (92.8%), mainly by the 20–24-year-olds. Although adolescent boys aged 15-19 year had heard of condoms, none of them were using it. About 28.6% reported using female condoms with their partners and about the same percentage reported their partners were using pills. At least 21.4% reported their partners used emergency contraceptives (EC). The least used methods were IUD and implants by their partners, and non-reported using the LARMs (female and Male sterilization) (See table 2).

**Table 2:** Current methods used by young people aged 15-24 years.

| FP Methods | Women |  | Men |  |
| --- | --- | --- | --- | --- |
|  | 15-19 (n=2) | 20-24 (n=29) | 15-19 (n=0) | 20-24 (n=14) |
| IUD | 0 (0.0) | 1 (3.2) | 0 (0.0) | 1 (7.1) |
| Injectables | 2 (6.5) | 18 (58.1) | 0 (0.0) | 1 (7.1) |
| Implants | 0 (0.0) | 13 (41.9) | 0 (0.0) | 2 (14.3) |
| Pills | 0 (0.0) | 2 (6.5) | 0 (0.0) | 4 (28.6) |
| Male condom | 0 (0.0) | 4 (12.9) | 0 (0.0) | 13 (92.8) |
| Female condoms | 0 (0.0) | 1 (3.2) | 0 (0.0) | 4 (28.6) |
| EC | 0 (0.0) | 1(3.2) | 0 (0.0) | 3 (21.4) |
| LAM | 0 (0.0) | 2(6.5) | 0 (0.0) | 1 (7.1) |

### 3.3 Multivariable analysis of contraceptive use

In the multivariable analysis, factors significantly associated with FP use for both females and males were age, education level, having children, living with biological parents, and having paid job. Being married was significant for women while listening to the radio and religion were significant for men (table 3).

**Table 3:** Factors associated with contraceptive use among youth aged 15-24 years.

| Socio demographic characteristics | Women |  | Men |  |
| --- | --- | --- | --- | --- |
|  | Unadjusted prevalence Rate (UPR) | Adjusted Prevalence Rate (APR) | Unadjusted prevalence Rate (UPR) | Adjusted Prevalence Rate (APR) |
| **Age** |  |  |  |  |
| 20-24 | 1.0 | 1.0 | 1.0 | 1.00 |
| 15-19 | 0.95 (0.75-1.20) *** | 0.70 (0.63-0.77) ** | 0.62 (0.48- 0.79) ** | 0.58 (0.47-0.72) *** |
| **No education\*** | 1.00 | 1.0 | 1.00 | 1.00 |
| Primary | 0.98 (0.81-1.18) *** | 0.73 (0.66-0.81) ** | 0.43 (0.34-0.56) | 0.50 (0.41-0.61) |
| Secondary+ | 0.92 (0.71-1.19) *** | 1.32 (1.15-1.52) *** | 0.98 (0.34-0.56) ** | 1.25 (0.50-0.77) * |
| **Married\*** | 1.0 | 1.0 | 1.00 | 1.0 |
| Separated | 1.10 (0.75-1.61) * | 1.81 (1.23-2.62) *** | 1.9 (0.42-8.65) | 0.68 (0.16-2.94) |
| Never Married | 1.08 (0.77-1.52) * | 3.09 (2.39-3.99) *** | 0.78 (0.44-1.38) | 1.00 (0.61-1.64) |
| **Children** |  |  |  |  |
| No children | 1.0 | 1.0 | 1.0 | 1.0 |
| Have children | 1.12(0.82-1.52) *** | 1.46 (1.18-1.81) *** | 6.8 (3.97-11.64) *** | 7.4 (4.74-11.69) *** |
| **Lives with** |  |  |  |  |
| Biological parents | 1.0 | 1.0 | 1.0 | 1.0 |
| Guardians | 0.93 (0.68-1.26) ** | 0.66 (0.57-0.77) *** | 0.11 (0.06-0.19) *** | 0.43 (0.32-0.59) *** |
| Older siblings | 0.77 (0.40-1.47) ** | 0.16 (0.08-0.30) *** | 0.19 (0.04-0.82) * | 0.12(0.03-0.54) ** |
| Own | 0.98 (0.77-1.25) *** | 0.33 (0.27-0.40) *** | 0.77 (0.05-0.12) *** | 0.13 (0.09-0.19) *** |
| Wife/husband | 1.00 (0.74-1.34) * | 0.92 (0.71-1.19) *** | 0.04 (0.01-0.12) *** | 0.21 (0.07- 0.63) ** |
| **No Job\*** | 1.0 | 1.0 | 1.0 |  |
| Farming/Agriculture | 1.06 (0.85-1031) | 2.17 (1.94-2.24) *** | 10.44 (7.30-14.94) *** | 5.5 (4.13-7.25) *** |
| Other | 0.98 (0.80-1.21) | 4.51(3.80-5.36) *** | 2.07 (1.60-2.68) *** | 1.6 (1.26-1.92) *** |
| **Religion** |  |  |  |  |
| Protestants | 1.0 |  | 1.0 |  |
| Catholic | 1.00 (0.83-1.19) |  | 0.24 (0.18-0.33) *** |  |
| Muslim | 0.90 (0.62-1.31) |  | 0.41 (0.29-0.58) *** |  |
| Other | 0.97 (0.52-1.81) |  | 0.40 (0.54-3.07) |  |
| **Listening to radio** |  |  |  |  |
| Almost every day | 1.00 | 1.00 | 1.00 | 1.0 |
| Once a week | 1.07 (0.87-1.32) * | 0.60 (0.50-0.70) | 0.62 (0.39-1.01) ** | 0.41 (0.26-0.64) *** |
| Less than once a week | 1.19 (0.83-1.69) | 1.12 (0.80-1.58) | 1.21(0.16-9.26) | 0.34 (0.04-2.51) |
| Not at all | 1.23 (1.00-1.51) | 2.46 (2.19-2.77) | 5,65 (4.08-7.84) *** | 2.41(1.89-3.07) *** |

\*\*\*p<0.001 \*\*p<0.01 \*p<0.05

Females and males aged 15-19 years had lower prevalence by 30% (APR=0.70, 95%CI: 0.63-0.77) and 42% (APR= 0.58, ci: 0.47-0.72) respectively compared to those aged 20-24 years. Females and males who were in or had attained secondary education had a higher prevalence of 32% (APR=1.32, 95%CI: 1.15-1.52) and 25% (APR= 1.25, 95% CI: 0.50-0.77) respectively compared to those with no education. Female and male youth with paid jobs were 4.5 times (APR=4.51, 95%CI: 3.80-5.36) and 1.6 times (APR=1.6, 95% CI: 1.26-1.92) more likely to use contraceptives respectively compared to those with no jobs.

The never married female youth were 3.1 times more likely to use FP than married female youth (APR=3.09, 95%CI: 2.39-3.99) and the prevalence of FP use was 46% higher among female youth who had biological children compared to those with no biological children (APR=1.46, 95%CI: 1.18-1.81) and men who had biological children were 7.4 times more likely to use contraceptives (APR=7.4, 95% CI: 4.74-11.69).

Male youth who were Catholics and Muslims had 76% (APR=0.24, 95%CI: 0.18-0.33) and 59% (APR=0.41, 95%CI: 0.29-0.58) lower prevalence rates respectively compared to Protestants. There was no significant difference in prevalence rate by religion for females.

### 3.4 Multivariable analysis for intention to use FP

Table 4 shows that the factors significantly associated with intention to use FP were age, education, marital status, living with biological parents, having paid job and farming, religion and listening to a radio while having children was significant only for males. The prevalence of intention to use FP for female youth aged 15-19 years was half (50%) that of 20–24-year-olds (APR=0.50, 95% CI: 0.44-0.56) while the intention was lower by 28% among 15–19-year-old males (APR= 0.72, 95%CI: 0.53-0.95).

**Table 4:** FP Factors associated with intention to us FP among youth aged 15-24 years.

| Socio demographic characteristics | Women |  | Men |  |
| --- | --- | --- | --- | --- |
|  | Un adjusted | Adjusted | Un adjusted | Adjusted |
|  | Prevalence Rate<br>(UPR) | Prevalence Rate<br>(APR) | Prevalence Rate<br>(UPR) | Prevalence Rate<br>(APR) |
| **Age** |  |  |  |  |
| 20-24 | 1.0 | 1.0 | 1.0 | 1.0 |
| 15-19 | 0.52 (0.46-0.58) *** | 0.50 (0.44-0.56) *** | 0.56 (0.38-0.58) ** | 0.72 (0.53-0.95) ** |
| **Education level** |  |  |  |  |
| No education | 1.0 | 1.0 | 1.0 | 1.0 |
| Primary | 0.62 (0.55-0.68) *** | 0.48 (0.43-0.58) *** | 0.43 (0.29-0.62) *** | 0.46 (0.36-0.59) *** |
| Secondary+ | 0.35 (0.19-0.64) *** | 0.22 (0.10-0.51) *** | 0.12 (0.07-0.20) *** | 0.15 (0.09-0.24) *** |
| **Married** | 1.0 | 1.0 | 1.0 | 1.0 |
| Separated | 0.73 (0.49-1.09) | 0.85 (0.54-1.32) | 0.35 (0.04-2.96) | 0.93 (0.12-7.35) |
| Never Married | 2.4 (1.90-2.96) *** | 2.03 (1.58-2.60) *** | 2.39 (0.95-5.96) | 0.56 (0.32-0.99) * |
| **Children** |  |  |  |  |
| Have children | 1.0 | 1.0 | 1.0 | 1.0 |
| No children | 0.89 (0.70-1.11) | 0.99 (0.80-1.23) | 3.70 (1.61-8.49) ** | 3.15 (1.92-5.15) *** |
| **Lives with** |  |  |  |  |
| Biological parents | 1.0 | 1.0 | 1.0 | 1.0 |
| Guardians | 0.52 (0.43-0.62) *** | 0.42 (0.34-0.57) *** | 0.32 (0.13-0.76) ** | 0.44 (0.28-0.69) *** |
| Older siblings | 0.19 (0.09-0.38) *** | 0.08 (0.02-0.33) *** | 0.08 (0.01-0.65) ** | 0.02 (0.03-0.16) *** |
| Own | 0.37 (0.30-0.46) *** | 0.29 (0.22-0.38) *** | 0.76 (0.43-1.34) | 0.12 (0.07-0.17) *** |
| Wife/husband | 0.66 (0.55-0.79) *** | 0.66 (0.53-0.81) *** | 0.96 (0.12-0.48) ** | 0.02 (0.01-0.11) *** |
| **No Job** | 1.0 | 1.0 | 1.0 | 1.0 |
| Farming/Agriculture | 1.75 (1.41-2.16) *** | 3.58 (3.15-4.07) *** | 1.01 (0.28-3.48) | 0.92 (0.30-2.81) |
| Other | 2.00 (1.56-2.47) *** | 5.75 (4.94-6.69) *** | 2.94 (2.02-4.31) *** | 2.25 (1.76-2.89) *** |
| **Religion** |  |  |  |  |
| protestants | 1.0 |  | 1.0 |  |
| Catholics | 0.89 (0.79-1.00) |  | 1.06 (0.00-2.06) |  |
| Moslems | 0.19 (0.12-0.27) *** |  | 0.06 (0.02-0.20) *** |  |
| Seventh Day Adventist | 4.90 (3.88-6.38) *** |  | 3.95 (1.50-10.34) *** |  |
| **Listening to radio** |  |  |  |  |
| Almost every day | 1.0 | 1.0 | 1.0 |  |
| Once a week | 0.74 (0.64-0.86) *** | 0.63 (0.53-0.76) *** | 0.79 (0.41-1.50) | 5.71 (3.47-9.41) *** |
| Less than once a week | 0.61 (0.43-0.88) *** | 0.52 (0.35-0.78) *** | 0.15 (0.02-1.19) | 1.64 (0.21-13.05) |
| Not at all | 1.79 (1.56-2.04) *** | 1.91 (1.65-2.22) | 9.22 (5.54-15.36) *** | 10.7 (6.97-16.65) *** |

\*\*\*p<0.001 \*\*p<0.01 \*p<0.05

The never married females were 2.03 times more likely to have intentions to use FP compared to married females (APR=2.03, 95% CI: 1.58-2.60) while never married males had lower prevalence by 44% (APR=0.56, 95% CI: 0.32-0.99). Female youth living with other relatives including guardians, siblings, their spouses, or those living on their own had lower prevalence rates of intention to use FP by 58%, 99.9%, 34% and 69% respectively compared to those who lived with their biological parents while a similar trend was observed for male youth with lower intention to use contraceptives by 66%, 99.9%, 88% and 99.9% (see table 4). Female and male youths who had a job were 5.7 times (APR=5.75, 95%CI: 4.94-6.69) and 2.3 times (APR=2.25, 95%CI: 1.76-2.89) respectively more likely to have intention to use FP compared to those with no jobs.

Female and male youths who were Muslim significantly had a lower prevalence by 81% (UPR= 0.19, CI: 0.12-0.27) and 99.9% lower prevalence of intention to use FP (APR =0.06 95% CI: 0.02-0.20) compared to protestants. However, among Seventh Day Adventist the intention was higher by 4.9 times (UPR= 4.9, 95% CI: 3.88-6.38) and 3.9 times UPR= 3.95 95% CI: 1.50-10.34) among females and males respectively compared to protestants.

## 3. Discussion

This study found low contraceptive use among the youth. About 41.6% (49.3% males and 32.6% females) had ever used a contraceptive and only 11.0% (16.3% women and 6.4% men) were currently using a contraceptive, mainly among 20–24-year-olds compared to those 15-19 years. None of the adolescent boys (15-19 years) were currently using any method and only 1.1% of adolescent girls (15-19 years) were using a method even though they were sexually active. Male condoms were the most used method by men (92.8%) while women mainly used injectables 55.2% and implants (44.8%). The intention to use contraceptives was high at 72.4% (84.0% males and 59.0% females). The 15–19-year-olds had a 28% lower prevalence of intention to use contraceptives compared to the 20–24-year-olds.

Our study was conducted in Karamoja region, and the results are similar to another study conducted in Uganda in general among adolescents on modern contraceptive prevalence that found low prevalence at 9.4% only (6). Young people face many challenges accessing and using contraceptive services (9). The challenges commonly reported by previous studies include stockouts of preferred methods, lack of knowledge on how contraceptives work (10, 11, 12), myths and misconceptions, fear of side effects, provider attitudes (13), long waiting time, lack of privacy and social cultural norms (14, 15). Another study conducted in Uganda indicated that youth complained of lack of privacy as staff sometimes shout out private information in front of other service users and everyone will know why you are at the clinic (16). The lower utilization could be attributed partly to social and cultural norms around contraceptives, and limited availability of adolescent friendly health services (17). Our study adds to this existing literature where we found young people preferred condoms to other methods since it’s easier to get and carry especially for male youth where they can use their pockets. With condoms you spend less time in facilities obtaining them. We found that those who had children were more likely to use contraceptives. This adds to the believe or myth that unless you have children you can use contraceptives and if you use these methods before having children, you can become barren.

At the same time, we found that the use of contraceptives varied significantly among female and male participants. Uptake increased with improved levels of education, supporting existing literature both in Uganda and elsewhere (18 & 19,). Education improves young people’s agency and empowers them to make informed decisions (20), while educated youth are more likely to have knowledge about the existing family planning services, side effects, and how methods work (21 &22).

The high intention to use contraceptives among the youth revealed in this study is like findings from numerous studies previously conducted both in Uganda and other parts of sub-Saharan Africa). The high intention among youth in Karamoja to use contraceptives could be associated with a desire to have fewer children that they are able to provide for (23). Other theories have suggested that the increasing desire to use modern family planning among the Karamojong people reflects a shift from reliance on traditional natural method of child spacing and is heavily shaped by the rapid changes in the structure and economy of this largely pastoral community.

Nomadic pastoralism, the traditional practice among the Karamojong, where men left their families behind and went about roaming with their livestock, for up to months, in search for pastures and water ensured that men did not have access to their wives, acting as natural system of spacing birth (24). However, with nomadic lifestyles no longer tenable for many this natural method of birth spacing that relied on men’s absence from home may no longer be effective “thus, our findings show that youth have strong desires to use FP but that current services and structural factors have not been able to adequately meet (or satisfy) young people’s desires to use FP.

The result that married female youth were less likely to use contraceptives than the never marrieds contradicts a study by Fatuma N et al (2022) in Wakiso and Kamuli districts in Uganda among adolescents where they found that those who were married were more likely to use modern contraceptives (19), but is consistent with findings from another study in DR Congo that found that married young women were less likely to use modern contraception (25). In the study area, women are expected to give birth immediately after getting married to prove their fertility and justify the dowry paid (26). The high value attached to children discourages contraceptive use, particularly in contexts where women’s gender identities and social status are tied to motherhood (27), while the never marrieds tended to report less use of contraceptives, our own qualitative study (Anonymized) suggests that they might be using contraceptives secretly, since sex in Karamoja is only encouraged after marriage, with most community members viewing sex outside marriage as shameful and against their cultural values (26).

Our study found that having paid job had positive relationship with contraceptive use and intention to use contraceptives. Low socio-economic status has been independently linked to adolescent pregnancy while contraceptive use is linked with high socio-economic status (28). It is also likely that those with jobs have higher level of education and more knowledgeable of where to get contraceptives, how to manage the side effects, and how the methods work than those who do not have jobs. It has been shown that the more educated and richer one is, the more likely such a person will use contraceptives (29). Having income may also facilitate young people’s access to contraceptives (30). Young people who have jobs can afford costs associated with contraceptive use such as transportation where studies have revealed that it hinders women from seeking contraceptive services (31).

The finding that youth who lived with their biological parents had higher prevalence of contraceptive use and intention to use is inconsistent with many studies that show that parents do not support adolescents’ and young people to use modern contraceptives (32, 33). A study in Arua city Uganda revealed that parents or guardians were not in support of adolescents using modern contraceptives. Lack of parental support was related to perceptions that modern contraceptives promote sexual promiscuity, fear that it causes infertility and that it is incompatible with cultural, religious, and moral norms (34). Other studies in Uganda have, however, reported that information from parents on contraceptives may be regarded highly by adolescents as these were considered as authorities who held their best interests at heart

(31,35,36).

## 4. Implications and recommendations for future research, policies and programs

One important implication is that this is one of very few studies done on FP use in Karamoja covering both female and male youth, a region with one of the lowest prevalence of FP in Uganda - so our findings provide important data for programs and policies to target and develop specific programs that can target youth with certain characteristics who want/intend to use FP in the region

We provided specific recommendations of what can work for both male and female youth and what can work for only male or female youth. We recommend that emphasis should be put on improving access to education for female youth in Karamoja. We found that contraceptive use increased with increasing level of education for females, so keeping girls in schools will address uptake but also delay in getting pregnant

Create employment for young people both males and females who are out of school. We found having paid jobs by both males and females had positive associations with contraceptive use.

Increasing focus of contraceptive use for adolescents (15-19 years) since they had high intention to use contraceptives, involve parents in adolescents and young people’s SRH since those youth who were living with their parents were more likely to use contraceptives. Lastly, having a strategy for involving men to improve contraceptive use among young people.

## 5. Strengths and limitations

Many studies on contraceptive use among young people, tend to focus on adolescent girls and young women and rarely analyze data from female and male youth separately. We believe that conducting this study among the sexually active youth 15-24 years and analyzing the similarities and differences in factors associated with contraceptive use and intention to use among male and female youth offers valuable insights for tackling contraceptive use with a gender lens.

Our study had some limitation and challenges implementing this study. The study had few youths using contraceptives and the results here are based on those small numbers. Karamoja is generally a pastoral community and used postpartum abstinence to space births. We did not measure postpartum abstinence because it is a traditional method, and we were looking at modern contraceptives. Contraceptive utilization is just picking up slowly in this area, so the few numbers of youth using contraceptives might have had an influence on the contraceptive use results.

## 6. Conclusions

The factors positively associated with intention to and use of contraceptive use in this study include age, education level, never married status, having paid jobs, having biological children, listening to a radio, and living with biological parents. The factors that were negatively associated were living with siblings, spouses and guardians, religion, and being married. Low contraceptive utilization among youth is likely to affect achieving Sustainable Development Goals (SGDs).

## Data Availability

All relevant data are within the manuscript and its Supporting Information files.

## 7. Declarations

### Ethics approval and consent to participate

This is part of the PhD study and was cleared by the Makerere University School of Public Health Research Ethics Committee and Uganda National Council of Science and Technology.

### Consent for Publication

Not applicable

### Availability of data and materials

*The data is available and can be shared if required*.

### Ethics statement

This study was conducted in accordance with the Helsinki Declaration. This was part of a PhD study that was approved by the Makerere University School of Public Health, Research Ethics Committee (MUSPH-REC), approval number (SPH-2022-294). The study was also approved by Uganda National Council of Science and Technology (UNSCT), a national approving body (Internal Review Board), The UNCST approval number for the study is HS2547ES. Districts where the study was conducted provided letters of approval. All respondents who participated in the study provided verbal informed consent before the interview could start. Respondents were assured of safety, confidentiality, voluntary participation and withdraw from the survey without any penalty. For participants who were younger than the age of 18 years, informed consent was obtained from their parents or guardians

## Authors contributions

Lillian Ojanduru conceptualized the study, led data collection and analysis, and prepared the initial draft of the manuscript. Nazarius. *M.* Tumwesigye, reviewed tools, provided guidance on methodology, data analysis, reviewed and approved manuscript. Godfrey Sui, and Justine Bukenya reviewed the tools, the draft manuscript, and approved the manuscript.

## 8. Acknowledgements

The authors would like to thank Makerere University for providing funds for data collection under the Research Innovations Fund (RIF), Round 4, track 2. We thank Dr Ether Janaina Spindler, for providing editorial and language review for the Manuscript We thank all our research participants for taking time to participate in the study.

